# ChatGPT Goes to Operating Room: Evaluating GPT-4 Performance and Its Potential in Surgical Education and Training in the Era of Large Language Models

**DOI:** 10.1101/2023.03.16.23287340

**Authors:** Namkee Oh, Gyu-Seong Choi, Woo Yong Lee

**Affiliations:** Department of Surgery, Samsung Medical Center, Sungkyunkwan University School of Medicine, Seoul, Korea

**Author notes:** **Corresponding author** Professor, Woo Yong Lee, M.D., Ph.D., Department of Surgery, Samsung Medical Center, Sungkyunkwan University School of Medicine, Address: 81 Irwon-ro, Gangnam-gu, Seoul, Republic of Korea 06351.

**Keywords:** Artificial intelligence, General surgery, Medical education, Continuous medical education

## Abstract

**Purpose:** This study aimed to assess the performance of ChatGPT, specifically the GPT-3.5 and GPT-4 models, in understanding complex surgical clinical information and its potential implications for surgical education and training.

**Methods:** The dataset comprised 280 questions from the Korean general surgery board exams conducted between 2020 and 2022. Both GPT-3.5 and GPT-4 models were evaluated, and their performances were compared using McNemar’s test.

**Results:** GPT-3.5 achieved an overall accuracy of 46.8%, while GPT-4 demonstrated a significant improvement with an overall accuracy of 76.4%, indicating a notable difference in performance between the models (P < 0.001). GPT-4 also exhibited consistent performance across all subspecialties, with accuracy rates ranging from 63.6% to 83.3%.

**Conclusion:** ChatGPT, particularly GPT-4, demonstrates a remarkable ability to understand complex surgical clinical information, achieving an accuracy rate of 76.4% on the Korean general surgery board exam. However, it is important to recognize the limitations of LLMs and ensure that they are used in conjunction with human expertise and judgment.

## INTRODUCTION

Significant advancements in large language model (LLM) technology have recently revolutionized the field of artificial intelligence (AI), with ChatGPT released by OpenAI in November 2022 standing out as a prime example [1]. ChatGPT has exhibited exceptional performance in evaluating knowledge related to fields such as medicine, law, and management, which have traditionally been considered to be the domain of experts. Notably, the system achieved high accuracy on the USMLE, the Bar exam, and the Wharton MBA final exam, even without fine-tuning the pre-trained model [2-5].

Surgical education and training demand a significant investment of time, with the process involving a combination of didactic learning, hands-on training, and supervised clinical experience [6]. During residency, surgical trainees work alongside experienced surgeons, gaining practical experience in patient care, surgery, and clinical decision-making.

Additionally, trainees engage in a series of didactic courses and conferences covering the principles of surgery, medical knowledge, and surgical techniques. Due to the comprehensive nature of surgical education and training, it can take more than a decade to become a skilled and competent surgeon. Given the time-intensive nature of surgical education and training, it is important to explore how emerging technologies, such as AI and LLMs, can augment the learning process [7].

This study aims to employ ChatGPT to evaluate the general surgery board exam in Korea and assess whether LLMs possess expert-level knowledge. Moreover, the study compared the performance of GPT-3.5 and GPT-4. By exploring the potential of LLMs in the context of surgical education and training, this study seeks to provide a foundation for future research on how these advancements can be effectively integrated into clinical education and practice, ultimately benefiting surgical residents, and practicing surgeons.

## METHODS

### General surgery board exam of Korea

The goal of surgical education and training is to develop the ability to actively evaluate the pathological conditions of surgical diseases and to acquire the surgical skills to treat traumatic, congenital, acquired, neoplastic, and infectious surgical diseases. To quantitatively evaluate this knowledge and skill set of surgical residents, a board certification exam is required after completion of their training, in order to become a board-certified general surgeon in Korea. The exam is composed of two parts: the first part is a 200-question multiple-choice test, and those who pass the first part are eligible to take the second part. The second part consists of questions based on high-resolution clinical images and surgical video clips. The questions are created and supervised by the Korean Surgical Society (KSS) and the Korean Academy of Medical Science (KAMS).

### Dataset for model testing

The actual board exam questions are held by KAMS, but due to limited access to the usage of these questions, we constructed our dataset by gathering questions recalled by examinees who took the actual exam. As the LLM cannot process visual information such as clinical images, radiology, and graphs, questions that included visual information were excluded from our dataset. All problems were manually inputted in their original Korean text. Finally, our dataset included a total of 280 questions from the first stage of the board exam in 2020, 2021, and 2022 (Figure 1. A).

**Figure 1.**
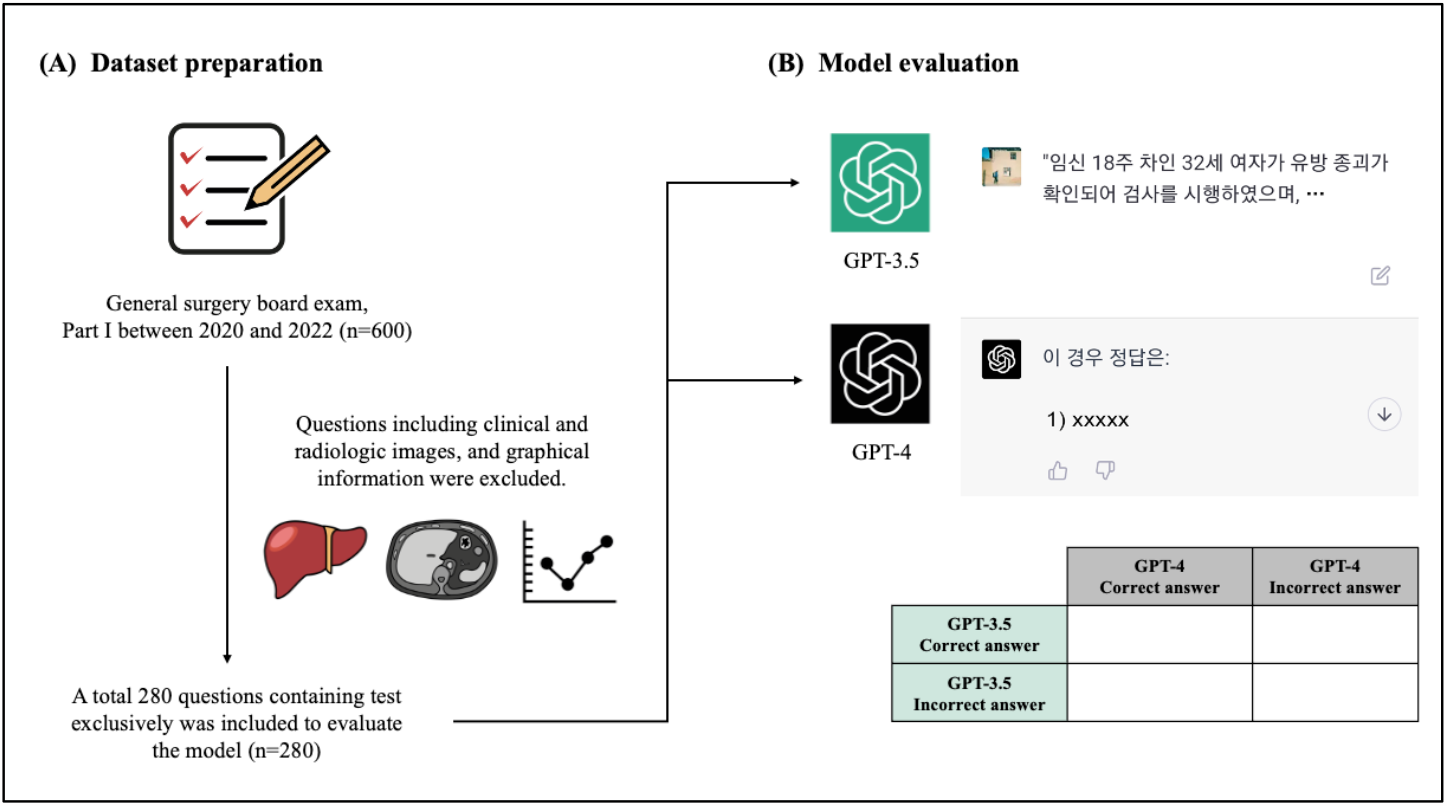
(A) Dataset preparation process for model evaluation, (B) How models were evaluated from ChatGPT website.

### Large language model and performance evaluation

In this study, we utilized the ChatGPT generative pre-trained transformer (GPT) language model developed by OpenAI to evaluate its performance on a dataset of questions. We performed model testing using both GPT-3.5 and GPT-4, with the former conducted from March 1st to March 3rd, 2023, and the latter scheduled for March 15th, 2023. To evaluate the model’s performance, we manually entered the questions into the ChatGPT website and compared the answers provided by the model to those provided by examinees (Figure 1. B).

### Statistical analysis

This study compared the performance of the GPT-3.5 and GPT-4 models with the McNemar’s test. A p-value less than a 0.05 would indicate a statistically significant difference between the performance of the GPT-3.5 and GPT-4.

### Ethical approval

The study did not involve human subjects and did not require institutional review board approval.

## RESULTS

The dataset used for model evaluation consisted of a total of 280 questions, which were classified into subspecialties and listed in order of frequency as follows: endocrine (16.8%), breast (16.1%), lower gastrointestinal (LGI, 14.3%), upper gastrointestinal (UGI, 13.2%), general (13.2%), pediatric (6.4%), hepatobiliary and pancreas (HBP, 6.1%), vascular (6.1%), transplantation (4.0%), and trauma and critical care(4.0%). (Figure 2)

**Figure 2.**
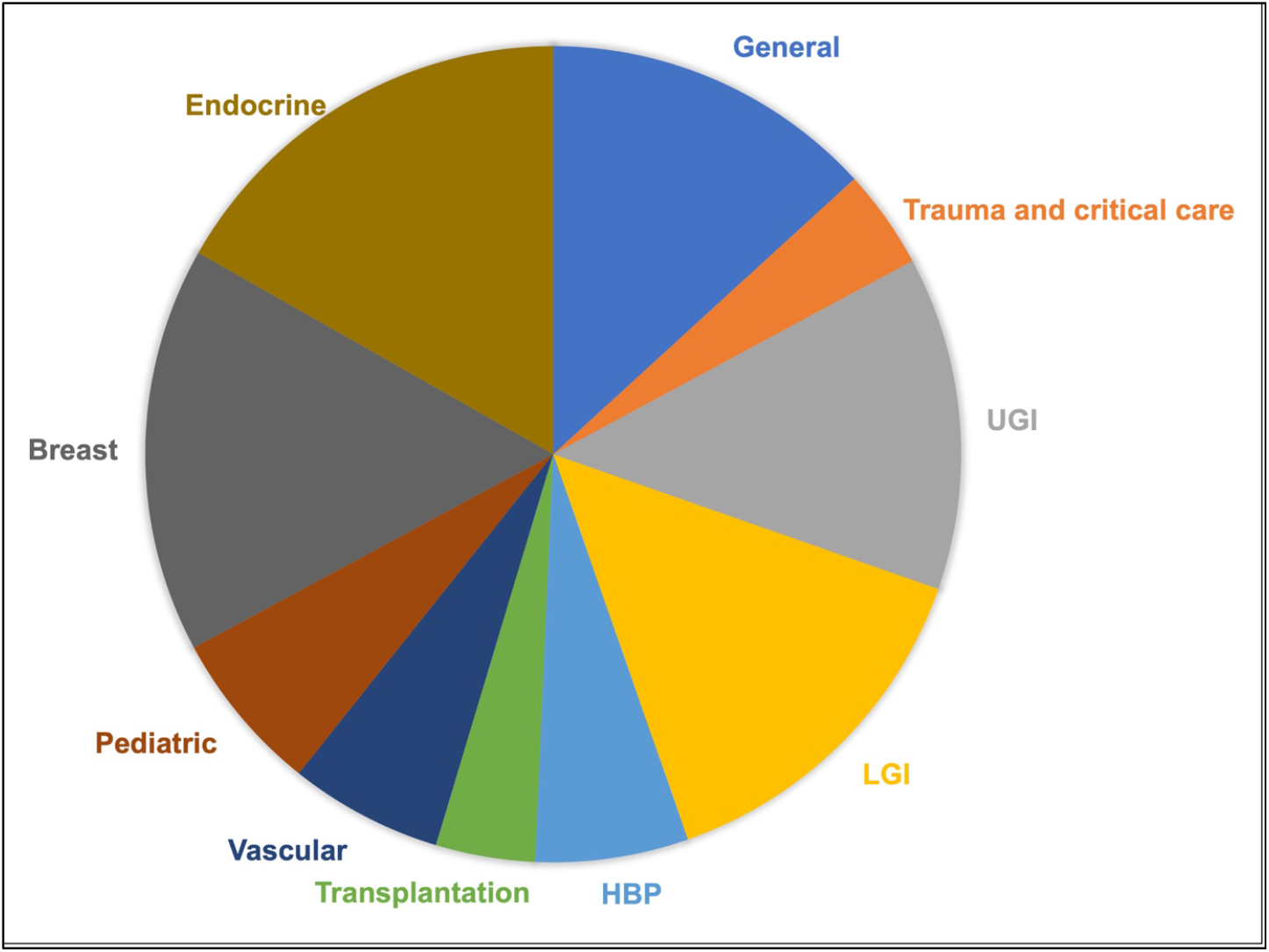
The dataset was composed of 280 questions, and it is classified into subspecialties in the field of general surgery.

A significant difference in performance was observed between the GPT-3.5 and GPT-4 models (P < 0.001). The GPT-3.5 model achieved an overall accuracy of 46.8%, providing correct answers for 131 out of the 280 questions. (Table 1) In terms of individual subspecialties, the model’s accuracy rates were as follows (sorted from highest to lowest): transplantation (72.7%), breast (62.2%), HBP (52.9%), general (48.6%), UGI (45.9%), trauma and critical care (45.5%), LGI (45.0%), endocrine (36.2%), pediatric (33.3%), and vascular (29.4%). In contrast, the GPT-4 model demonstrated a substantial improvement in overall accuracy, attaining a rate of 76.4% by providing correct answers for 214 out of the 280 questions. The accuracy rates for each subspecialty were as follows: pediatric (83.3%), breast (82.2%), UGI (81.1%), endocrine (78.7%), general (75.7%), transplantation (72.7%), LGI (72.5%), vascular (70.6%), HBP (64.7%), and trauma and critical care (63.6%). (Figure 3)

**Table 1.** Comparison table for the accuracy of GPT-3.5 and GPT-4.

| Table 1. Comparison table for the accuracy of GPT-3.5 and GPT-4. |  |  |
| --- | --- | --- |
|  | GPT-3.5 | GPT-4 |
| Correct answer | 131 | 214 |
| Incorrect answer | 149 | 66 |
| Accuracy | 46.8% (131/280) | 76.4% (214/66) |

GPT: generative pre-trained transformer

**Figure 3.**
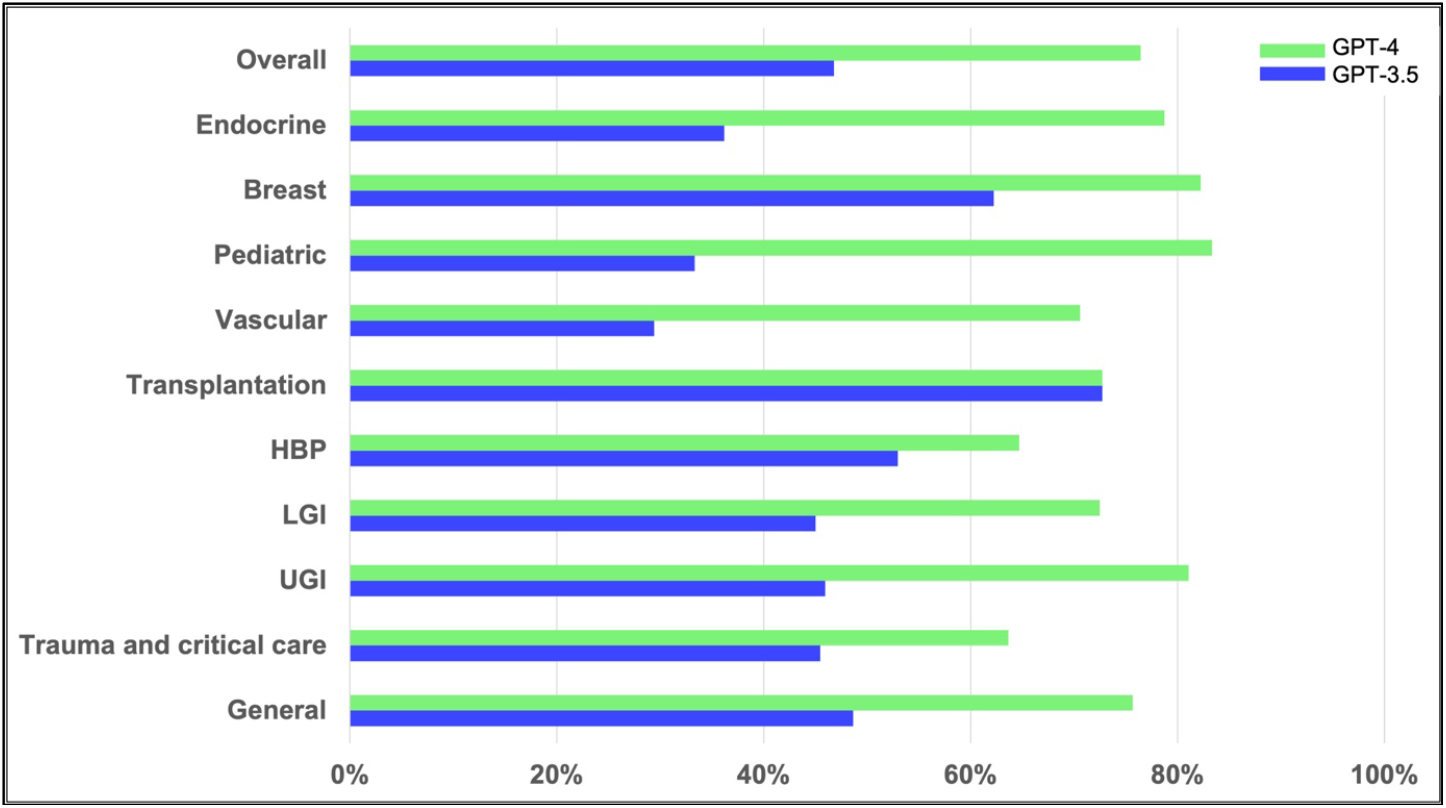
Comparison of the performance of GPT-4 and GPT-3.5 with overall accuracy and accuracies according to its subspecialties.

## DISCUSSION

The primary objective of this study was to conduct a quantitative assessment of ChatGPT’s ability to comprehend complex surgical clinical information and to explore the potential implications of LLM technology for surgical education and training. Specifically, we tested the performance of ChatGPT using questions from the Korean general surgery board exam and observed that the model achieved an accuracy of 76.4% with GPT-4 and 46.8% with GPT-3.5. Remarkably, this accuracy was achieved without fine-tuning the model and by using prompts in Korean language exclusively, thus highlighting the significance of our findings.

The comparative analysis revealed a notable improvement in GPT-4’s performance compared to GPT-3.5 model across all subspecialties. GPT-4 not only exhibited a higher overall accuracy rate but also demonstrated more consistent performance in each subspecialty, with accuracy rates ranging from 63.6% to 83.3%. However, for 18 questions, GPT-3.5 provided the correct answer while GPT-4 did not (Table 2). It is unclear why GPT-4 gave incorrect answers for these questions despite the overall increase in accuracy.

**Table 2.** A 2 by 2 contingency table summarizing performance of GPT-3.5 and GPT-4.

|  | GPT-4 correct answer | GPT-4 incorrect answer |  |
| --- | --- | --- | --- |
| GPT-3.5 correct answer | 113 | 18 * | 131 |
| GPT-3.5 incorrect answer | 101 | 48 | 149 |

GPT: generative pretrained transformer

Pinpointing the exact reason for this discrepancy is challenging. Differences in training data, model architecture, or other factors could have contributed to the variation in the performance between the two versions.

The authors kindly recommend that the surgeon’s society proactively adapts and utilizes these technological advancements to enhance patient safety and improve the quality of surgical care. In the context of surgical education, it is crucial to transition from the traditional rote learning approach to a method that emphasizes problem definition in specific clinical situations and the acquisition of relevant clinical information for problem resolution. LLMs serve as generative AI models, providing answers to given problems. Consequently, the quality of the answers relies on the questions posed [8]. Surgeons must conduct thorough history taking and physical examinations to accurately define the problems they face. By providing LLMs with comprehensive summaries of patients’ chief complaints, present illnesses, and physical examinations, the models have the potential to assist in decision-making regarding diagnostic tests and treatment options in certain clinical situations. However, it is essential for medical professionals to remember that LLMs should not replace the fundamentals of patient care, which include maintaining close connections with patients and actively listening to their concerns [9].

Moreover, active surgeons who completed their training over a decade ago may find LLMs helpful for continuous medical education (CME). Accessing new knowledge may be challenging for them due to the time elapsed since their training, potentially leading to outdated management practices. While numerous surgical societies offer CME programs, altering ingrained routines in clinical practice can be difficult. By utilizing an up-to-date LLM as a supplementary resource in their decision-making process, surgeons may have an additional means to stay informed and strive for evidence-based care in their patient management [10].

In medicine, decision-making has a profound impact on patient safety, demanding a higher level of accuracy and a conservative approach to change compared to other fields. Although GPT-4 achieved a 76.4% accuracy rate on the Korean surgical board exam, it is important to remember that LLMs are generative models, often reffered to as “stochastic parrots” [11].

Instead of providing strictly accurate information, they generate responses based on the probability of the most appropriate words given the data they have been trained on.

Consequently, the current level of accuracy is not yet sufficient for immediate clinical application in patient care.

However, it is noteworthy that a service released less than six months ago exhibits such remarkable performance, and ChatGPT is only one example of LLMs. Recently, Microsoft released BioGPT, an LLM trained on PubMed literature, and Meta introduced LLaMA, an LLM with an accessible API for open innovation and fine-tuning [12, 13]. Based on these trends, we can anticipate future LLMs to be trained on an even larger and more diverse set of medical information, providing specialized knowledge in the medical field. In addition, the GPT-4 framework itself is capable of processing and analyzing visual information, including images and videos [14]. This capability raises the possibility that, in the future, the performance of GPT-4 could be evaluated on datasets containing clinical photos and surgical videos. Such advancements would further enhance the applicability of GPT-4 in surgical fields, broadening its utility beyond text-based tasks and offering a more comprehensive understanding of complex clinical scenarios assisting professionals in their decision-making processes and contributing to improved patient care.

The limitations of this study include the fact that the dataset was compiled using questions recalled by examinees, which may not accurately represent the full set of actual board exam questions due to restricted access. Another limitation is the exclusion of visual information. Since the models used in the study are unable to process visual information, such as clinical images, radiology, and graphs, questions containing visual components were excluded from the dataset. As a result, we cannot determine whether ChatGPT would pass or fail the board exam based on these limitations. Despite these constraints, this study holds significance as it confirms the ability of LLMs to analyze surgical clinical information and make appropriate clinical decisions.

## CONCLUSION

ChatGPT, particularly GPT-4, demonstrates a remarkable ability to understand complex surgical clinical information, achieving an accuracy rate of 76.4% on the Korean general surgery board exam. However, it is important to recognize the limitations of LLMs and ensure that they are used in conjunction with human expertise and judgment.

## Data Availability

All data produced in the present study are available upon reasonable request to the authors

## Funding

This study was supported by Future Medicine 2030 Project of the Samsung Medical Center [SMX1230771].

The authors would like to thank Da Hyun Lee, an audiovisual engineer at Samsung Medical Information & Medical Services, for designing Figure 1 for this work.

## Notes

### Competing Interest Statement

The authors have declared no competing interest.

### Summary of Updates

The title and manuscript are revised during the peer review process to emphasize the potential implications of LLMs in surgical education and training, without overgeneralizing the findings. We have also clarified that the current study demonstrates the potential of LLMs in surgical education and training rather than making definitive conclusions.

